# The Easter and Passover Blip in New York City: How exceptions can cause detrimental effects in pandemic times

**DOI:** 10.1101/2020.04.14.20065300

**Authors:** Maximilian Vierlboeck, Roshanak R. Nilchiani, Christine M. Edwards

## Abstract

When it comes to pandemics such as the currently present COVID-19 [1], various issues and problems arise for infrastructures and institutions. Due to possible extreme effects, such as hospitals potentially running out of beds or medical equipment, it is essential to lower the infection rate to create enough space to attend to the affected people and allow enough time for a vaccine to be developed. Unfortunately, this requires that measures put into place are upheld long enough to reduce the infection rate sufficiently.

In this paper, we describe research simulating the influences of the contact rate on the spread of the pandemic using New York City as an example (Section IV) and especially already observed effects of contact rate increases during holidays [2-4] (Section V). In multiple simulations scenarios for Passover and Easter holidays, we evaluated 25%, 50%, 75%, and 100% temporary increases in contact rates using a scenario close to the currently reported numbers as reference and contact rates based on bioterrorism research as a “normal” baseline for NYC.

The first general finding from the simulations is that singular events of increased visits/contacts amplify each other disproportionately if they are happening in close proximity (time intervals) together. The second general observation was that contact rate spikes leave a permanently increased and devastating infection rate behind, even after the contact rate returns to the reduced one. In case of a temporary sustained increase of contact rate for just three days in a row, the aftermath results in an increase of infection rate up to 40%, which causes double the fatalities in the long run.

In numbers, given that increases of 25% and 50% seem to be most likely given the data seen in Germany for the Easter weekend for example [2, 3], our simulations show the following increases (compared to the realistic reference run): for a temporary 25% surge in contact rate, the total cases grew by 215,880, the maximum of required hospitalizations over time increased to 63,063, and the total fatalities climbed by 8,844 accumulated over 90 days. As for the 50% surge, we saw the total number of cases rise by 461,090, the maximum number of required hospitalizations increase to 79,733, and the total number of fatalities climb by 19,125 over 90 days in NYC.

All in all, we conclude that even very short, temporary increases in contact rates can have disproportionate effects and result in unrecoverable phenomena that can hardly be reversed or managed later. The numbers show possible phenomena before they might develop effects in reality. This is important because phenomena such as the described blip can impact the hospitals in reality. Therefore, we warn that a wave of infections due to increased contact rates during Passover/Easter might come as a result!

## I. Introduction, Situation, Problem

> *“A Pandemic Is the Worldwide Spread of a New Disease”* ^*[5]*^

The above mentioned definition by the WHO describes the current global situation in regard to the virus named COVID-19, that emerged world wide in the past months. As of the writing of this paper (April 13), there are 1,848,439 confirmed COVID-19 cases world wide and 117,217 confirmed deaths in over 200 countries [1]. The virus is confirmed to be transmissible from human to human [6, 7] and has constantly been spreading due to contact between individuals.

The problem with the spread is though, that while it seems like a simple mathematical model, it is dynamic complex system which does not necessarily behave in a linear way. Thus, predictions can be difficult and the actual behavior of the whole system, and therefore the outcome such as fatalities and infrastructure strain, is hard to evaluate. One way to conduct such evaluations is to design a representative model which simulates and mimics the real world phenomena as close as possible. With such a model, certain parameters and influences can be assessed by modifying the model and observing its reaction, which is what this paper is about.

Due to the importance of the above described transmission from human to human and the involved contact, the research presented in the following paragraphs took a look at the effective contact rates between humans in a theoretical dynamic simulation using New York City as an example, in order to determine what factors play what role and how certain influences interact. Therefore, various simulations and scenarios were assessed in order to discovered different behaviors and potential emergent phenomena based on and dependent on different factors.

The second section will describe the research methodology, the model utilized for the simulations, and how the specific simulations were conducted. Section III describes the assumptions that were made in order to design and set up the model as well as the involved parameter as a result. Section IV and V then demonstrate and discuss scenarios possible and likely in order to show the behavior of the system and certain emergent phenomena. Lastly, Section VI will summarize and discuss the outcomes and also give an outlook how research might continue.

## II. Model and Methodology

When looking at models for the spread of diseases, SIR models present a simple and easy to adapt starting point for such situations. SIR stands for “susceptible–infective-removed” and was first proposed by Kermack and McKendrick in 1927 [8]. The model is described as a differential system in which multiple factors depend on each other to determine the behavior of the three levels S, I, and R. The equations herein were as follows [also see 9]:

(I) Susceptible Population:

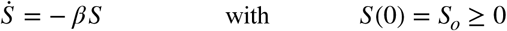

(II) Infectious Population:

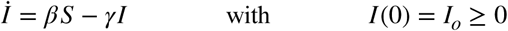

(III) Removed Population:

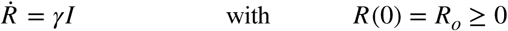

so that *S*(*t*)+*I*(*t*)+*R*(*t*) =*N* and 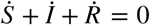

With these equations, a simulation system was be derived that models the current situation of the COVID-19 spread in a simplified way. Since the aforementioned infrastructure and hospital strain was of importance for the Coronavirus pandemic, the model was modified to include time delays due to incubation and a portion of the infected people who would not go directly from “infected” to “removed” and rather move to hospitalization. From hospitalization then there were two options, either a delayed demise of the individual, or a delayed recovery, which adds the individual back to R. These additions modify the equations above as follows and add equations (VII) and (VIII):

(IV) Susceptible Population:

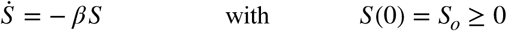

(V) Infectious Population:

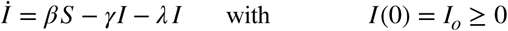

(VI) Removed Population (delayed):

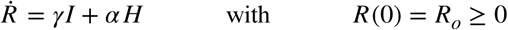

(VII) Hospitalized Population (delayed):

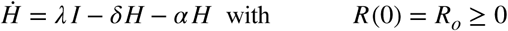

(VIII) Deceased Population (delayed):

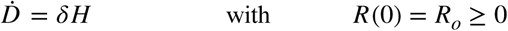

so that *S*(*t*)+*I*(*t*)+*R*(*t*)+*H*(*t*)+*D*(*t*)=*N* and 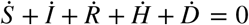

The simulation model based on these parameters was setup in Vensim [10] with time and calculation steps of one day. A flowchart of the model is depicted in Figure 1 on the right.

**Figure 1.**
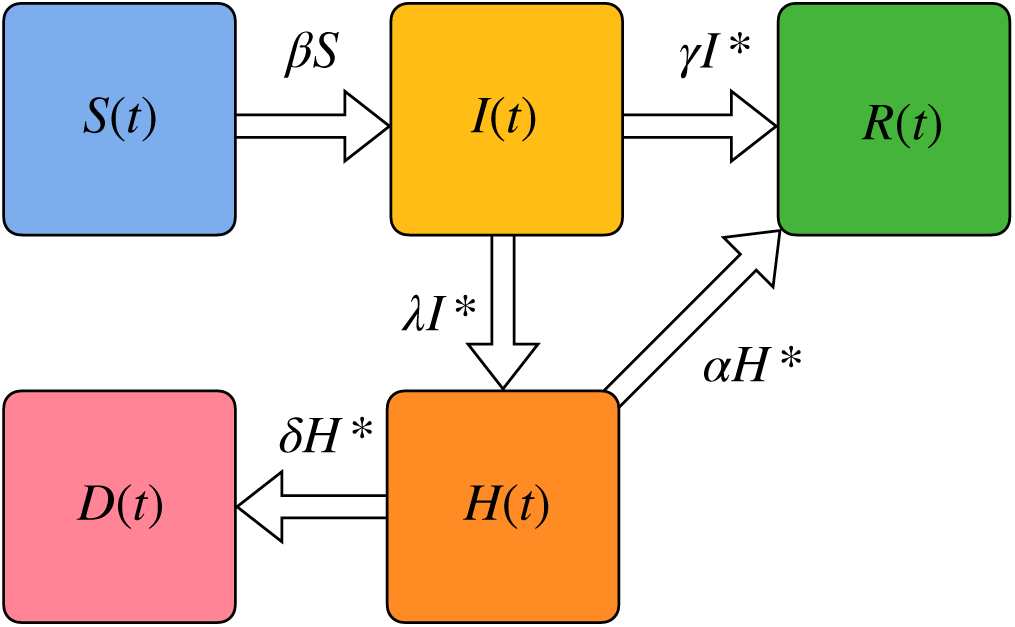
Simulation Flowchart (* marks delay impacts)

Based on the equations and the structure shown in Figure 1, the model was designed in order to allow for a flexible adjustment of the parameters, which will be described in Section 3. With the model then, the chosen research methodology was applied as described by Maria [11]. Herein, after the above described problem definition in a first step, the parameters of the model were set to yield an adequate and verifiable outcome. Such a verification was conducted by comparing the model results to real world data that was reported during the current pandemic.

Wirth the set parameters (also see next section), multiple scenarios were simulated and examined based on various conditions that were chosen, always derived from real and current circumstances. These scenarios will be described in the fourth and fifth section. The outcomes were compared as far as the different levels of the simulations components go. For example, the infection rates and total cases could be compared to determine the speed of the spread and therefore the rise of the total case number over time. Another option is the comparison and evaluation of fatality numbers and hospital strain over time to assess how different scenarios effect the end results and possibly discover potential shortages at certain times.

These scenarios then allowed for a general evaluation and also the discovery of the main focus of this paper, the phenomenon we called the “Easter Blip” (Section V). Based on the results, predictions of possible behaviors of the current pandemic were deduced to potentially support governing and regulating decision in order to avoid and mitigated unwanted situations such as high fatality numbers or collapse of medical support for example. The next section will describe the assumptions the model was based upon to allow for simulations that mimic the current real world behavior as far as feasible.

## III. Assumptions and Parameter

In order to design a model that could mimic and simulate the real world pandemic, the factors, described in the equations (IV) through (VIII) above, had to be set so that the simulation results would be in accordance with real world situations and data. Therefore, this section will outline the assumptions that were made to achieve the accordance. Hence, the following sub-sections will describe one parameter each based on New York City (NYC) in 2020, with a population of 8,398,748 people [12].

### The Parameter β - the Infection Rate

The infection rate of the model, which describes at what rate the susceptible population will be infected, was defined depending on two factors: infectivity (i) and effective contact rate (c). These two factors together with the infectious population (I) and the susceptible population (S) allow the calculation of the infection rate according to the following formula:

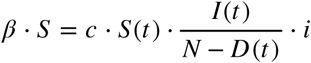

The infectivity (i) was defined as a constant based on the likelihood of infection when people interact and hence was derived from various sources and set to 5% [13, 14] due to the higher population density of NYC compared to the locations of the source data. The constant infectivity allowed a modulation and adjustment of the infection rate based on the second component, the effective contact rate. This rate was furthermore used to model and simulate real word behavior as circumstances like social distancing for example impact the effective contact rate of the population and therefore were ideal to be modeled this way. For the general magnitude of the effective contact rate, the amount of average contacts of people per day in NYC was researched in order to enable a realistic starting point without any measures such as social distancing.

Based on literature sources, the researched contact rate in NYC ranged from 5 for people who do not use the subway up to at least 10 for people who do utilize the subway [15]. Since this data was obtained and measured in 2003 and the population of NYC increased by 5% since then, this would yield contact rates of 5.25 and 12.5 today. Given that the number of subway users in NYC is higher than in any other city in the United States [16], it was assumed that 70% of the NYC population take the subway on a daily basis and therefore are more active and effectively have more contact, also through surfaces. Together with the number of contacts for non-subway users, this would yield an average effective contact rate without restrictions or social distancing of 10.325.

All in all, the infection rate therefore was defined by the following equation:

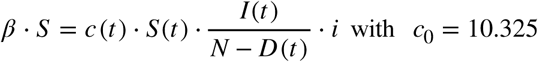

### The Parameters γ and λ - the Recovery and Hospitalization Rate

The parameters for the hospitalization and recovery rate were assumed to be directly connected as an infected person would either recover or be hospitalized (see Figure 1). Therefore, the recovery rate was exactly the opposite portion of the hospitalization rate, yielding

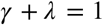

Since the numbers of hospitalizations strongly vary by age group and therefore depend on demographics, an average hospitalization rate was calculated based on official data by the City of New York [17] to allow for the use of a constant. The resulting probability was 0.27 for hospitalizations and thus 0.73 for recovery.

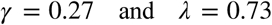

### The Parameters α and δ - the Hospital Recovery and Mortality

Similar to the last sub-section, the parameters for the hospital recovery and death rate were also assumed directly connected as a hospitalized person would either recover or decease. Therefore the hospital recovery rate was exactly the opposite portion of the death rate, yielding

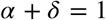

Since the death rate for people already hospitalized is much higher than the death rate of the virus in general, it was calculated based on the number of confirmed deaths and hospitalizations also provided by the City of New York [17], which resulted in a death rate of 0.223 and hence a hospital recovery rate of 0.777.

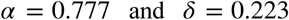

### Final Assumption - Unknown Numbers

The first positive COVID-19 case was reported in New York City on March 1^st^. Unfortunately, this is only the first confirmed positive case and not necessarily or likely the first case in general. Throughout the spread of the virus, only cases tested positive were reported and therefore a lack of people who carry the virus, but are not aware, has to continuously be assumed. This is further exacerbated by the fact that it is possible to carry the virus without ever showing symptoms [see 18]. Thus, the number of COVID-19 cases resulting from asimulation has to be way higher than what the real data represents. Actual numbers and estimation for the unknown numbers are hard to find and estimations range from over 70 percent unknown cases [19] to ten times the confirmed number or more [20]. Therefore, the number of unknown cases in the model was adjusted so that the model aligned from March 1^st^ to March 20^th^ with the reported real time data. In order to achieve this, the model was set to 15 infections at the time of the first reported case. This lead to a realistic outcome of the simulation and also served as verification of the design as the fatality rate and the case numbers correlated with the data when taken into consideration the unknown cases.

With these settings and parameters, the scenarios for the simulation could be run and evaluated. Since the measures and regulations that were put into place are hard to quantify, the first scenarios will address the effects of such measures and show how they could have affected the numbers. Then, the ensuing scenarios will evaluate possible future occurrences and possibilities. The following fourth section will cover these scenarios and therein discuss the general effects of the variable in the simulation, the effective contact rate. The fifth section then will discuss and show a possible and presumably likely phenomenon that could await in the near future, including its implications.

## IV. baseline Scenarios and General Effects

As described above, the first baseline scenario to assess is to figure out what trajectory the real world data most likely followed in order to understand what the measures that were put into place changed and how they affected the model. As mentioned, the variable to be manipulated will be the effective contact rate which directly affects the infection rate.

### Scenario 1 - Immediate Social Distancing and Bans Policy

The first measure that was put in place in NYC was social distancing and the closing of certain institutions and stores to be implemented immediately. This was accompanied by companies moving employees to work from home or stopping work all together. Official orders for example went into place on March 16^th^ and March 21^st^ after the national emergency was declared on March 13^th^ [see 21]. Therefore, the scenario below was constructed for the simulation to evaluate the effects. In a first run, the two dates were utilized to introduce step reductions in the effective contact rate of various heights and the effects on the infection rate were compared in form of a graph. Figure 2 and 3 below show the outcome for a period of 90 days, which corresponds to the time from March 1 through the End of May.

**Figure 2.**
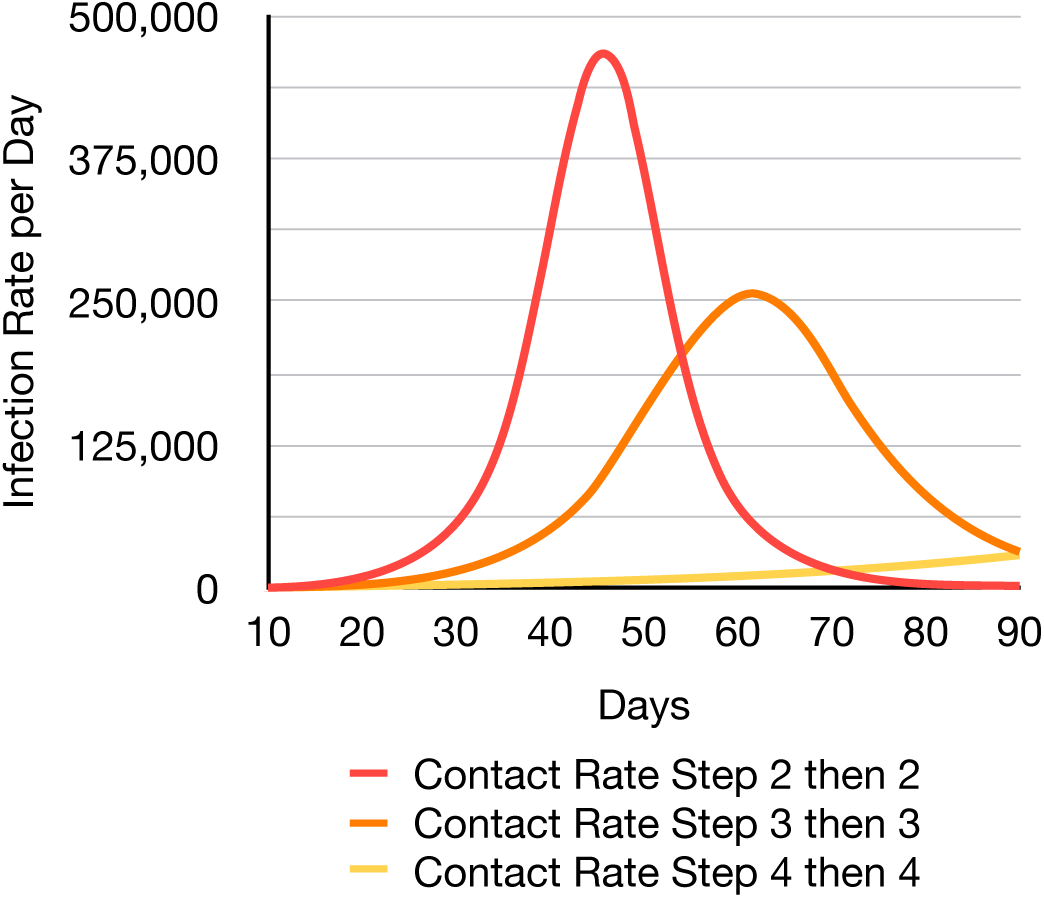
Scenario 1: Infection Rate Over Time

**Figure 3.**
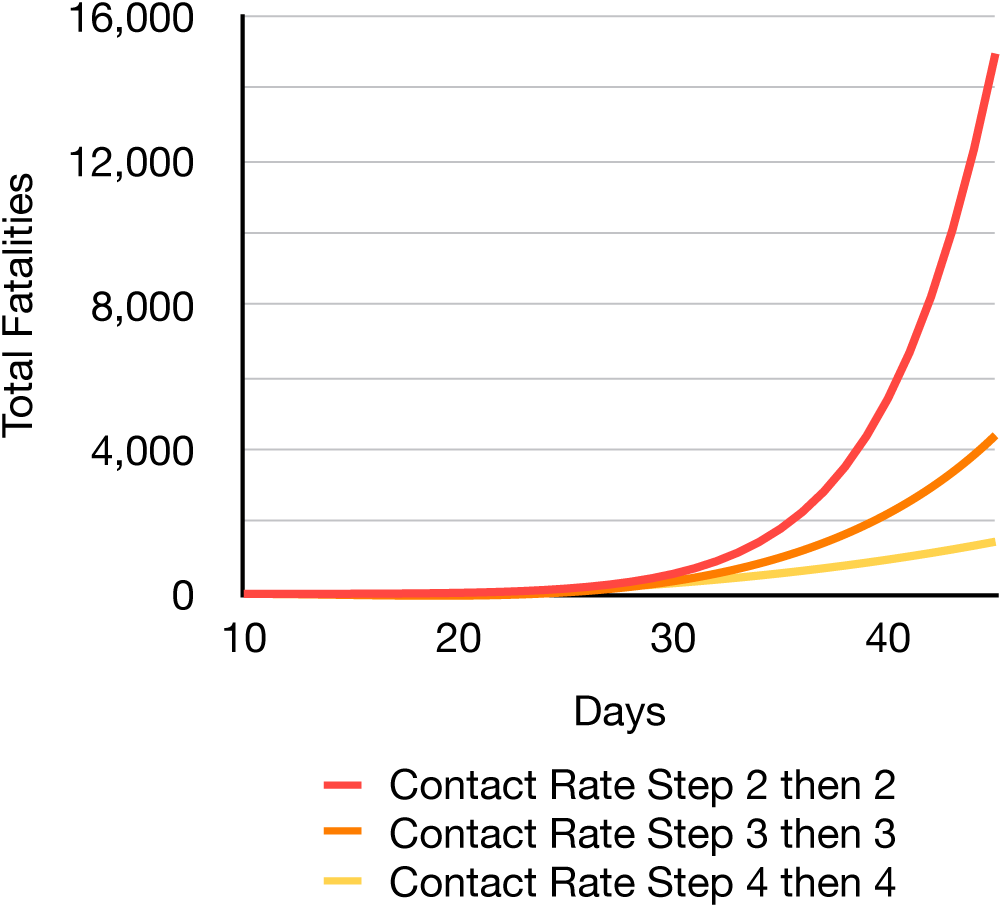
Scenario 1: Fatalities Over Time

Looking at the outcome, we see that the different effective contact rate steps flatten the curve significantly and stretch out the infections. Looking at the data as of April 10, which would correspond to the day 41 in the simulation, the fatality count in NYC was 5,429, which corresponds to the run above that included two steps with a reduction of 2 each time.

Unfortunately, as Figure 2 and 3 show, this version of Scenario 1 does not do well as far as the reduction of the infection rate goes over time and the fatalities keep increasing exponentially despite the measures. This is due to the fact that the reductions are not significant enough overall to have a helpful impact. Furthermore, such a scenario, while plausible and possible, is not realistic since measures put in place do not go into effect at once and everyone adheres to them immediately at the time they go into effect. Therefore, a continuous reduction is more realistic, which is why such a scenario will be presented and evaluated in Scenario 2.

### Scenario 2 : Gradual Social Distancing and Bans Policy

The second scenario, as above alluded to, will evaluate the effects of gradually reduced effective contact rates over a number of days. This way, the rates decrease over time until they reach certain events or a limit, which is more realistic since people adjust to new circumstances and in this case regulations gradually over time. Thus, the starting point of the regulations mentioned in the previous scenario was used to introduce effective contact rate reductions with a delay of one day. For example, the blue line in Figure 4 indicates a reduction of 1 for the effective contact rate after day 16 for 9 consecutive days until the rate reaches 1.325.

**Figure 4.**
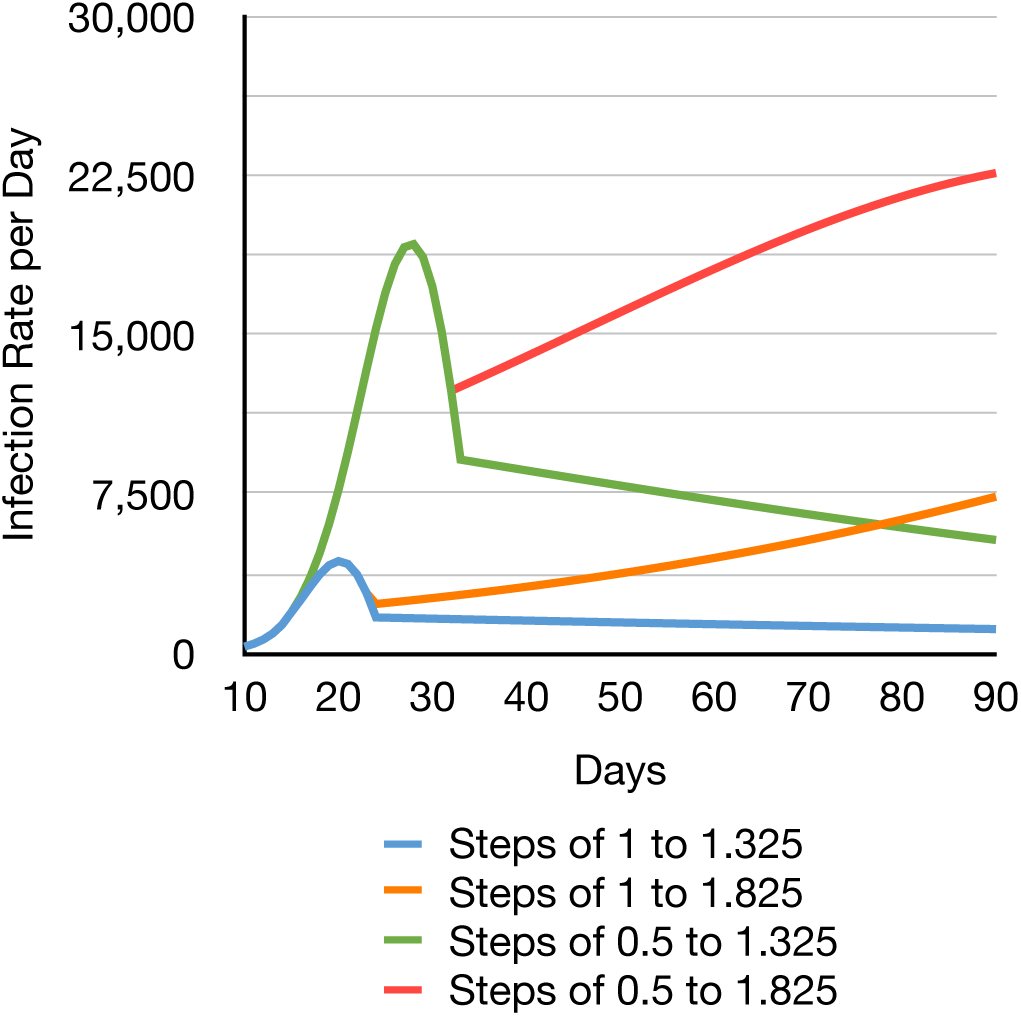
Scenario 2: Infection Rate Over Time

With this data, we can see that the run with the steps of 0.5 down to 1.825 is closest to reality and approaches the fatality number currently reported. Furthermore, we see that the gradual reduction of the effective contact rate leads to a peak in the infection rate which then introduces a downswing and successive upswing albeit the latter with a lower gradient of the Infection Rate over time. Therefore, we can say that the gradual reduction of the effective contact rate is an effective measure to control the epidemic and can even hedge the upswing of the virus spread, as shown by Figure 4.

## V. The Easter and Passover Blip

So far, we have looked at baseline scenarios which behave the same way over time and have changes that are linear or follow a gradient. Unfortunately, this is not at all the case in reality, as singular or short term relaxation in rules or temporarily making exceptions can cause major changes to the effective contact rate for a brief period of time. Such events can be short ones that increase the effective contact rate momentarily, but also longer time periods that show an increase or decrease, such as seasons, for example [see 22]. Since the increases are more critical than the decreases, we want to take a look at them in this section.

At the time of this writing (April 12, 2020), Easter is happening and during these times, various other religious holidays have happened or are coming up in the near future. During such holidays, people tend to congregate, visit religious gatherings such as masses, and visit family members. After a prolonged period of solitude, the perceived need and yearning for such close contacts increases understandably and there have already been reports of planned gatherings [4], measured significantly increased mobility in Germany [2, 3], and people (including two of the authors) have witnessed Good Friday gatherings at homes in New Jersey and New York, for example.

These phenomena raise the question what could happen if people are giving in to their yearnings and defy recommendations and regulations. Hence, this section will look at possibilities in two scenarios to estimate the implications of such defiance in order to enable a prediction regarding the outcome if the cause cannot be prevented. Scenario 3 will assess the possibility of increased effective contact rates on separate occasions and Scenario 4 will assess short periods of increases. As a basis for the scenarios, the trajectory closest to reality of Scenario 3 will be utilized.

### Scenario 3: Isolated Increases in Effective Contact Rate

To utilize a real life example we simulated the Run from Scenario 2 with the steps of 0.5 down to 1.325 and implemented two short increases in effective contact rate for Good Friday and Easter Sunday. In order to simulate various severities of increases, four runs were conducted with increments of 25% yielding the last run as a return all the way back to the effective contact rate c_0_ of 10.325. The results are depicted in Figure 6 through 9 on the next pages and discussed hereinafter.

**Figure 5.**
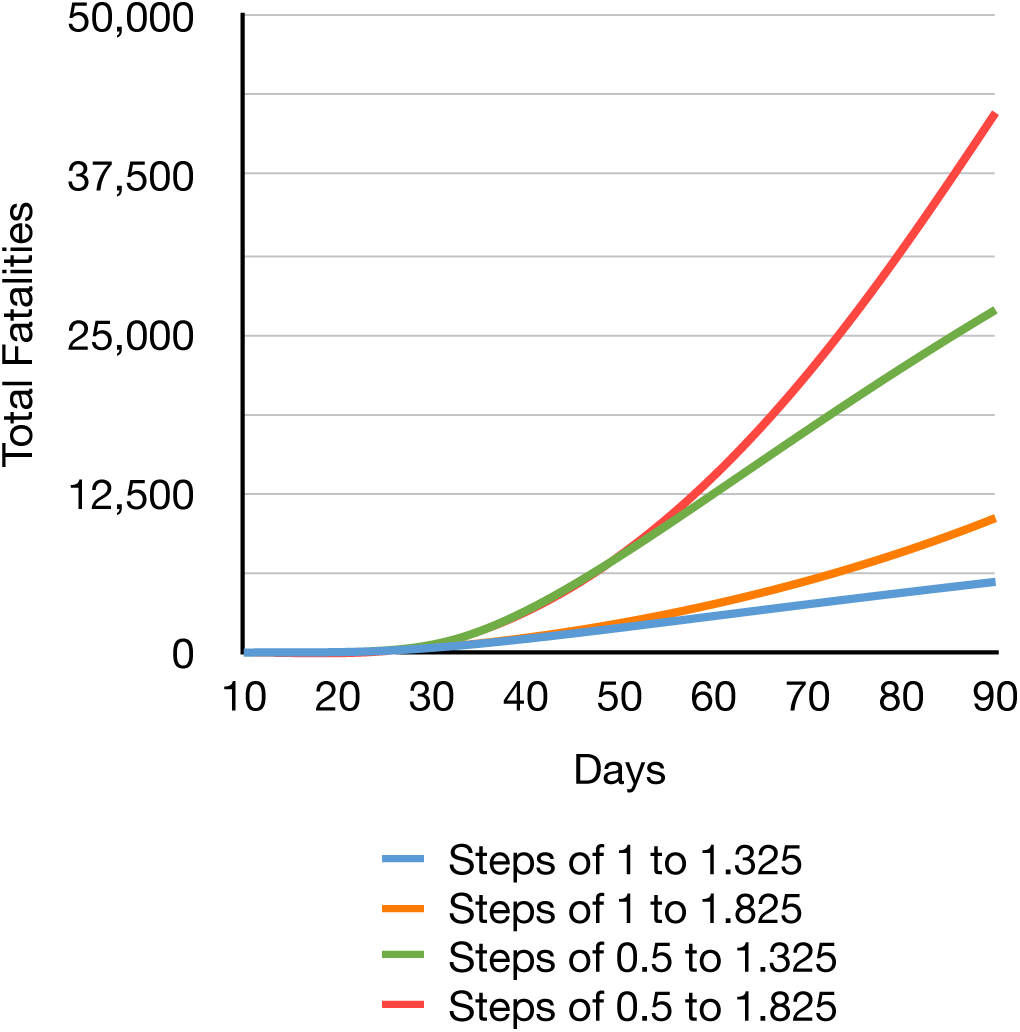
Scenario 2: Total Fatalities over Time

**Figure 6.**
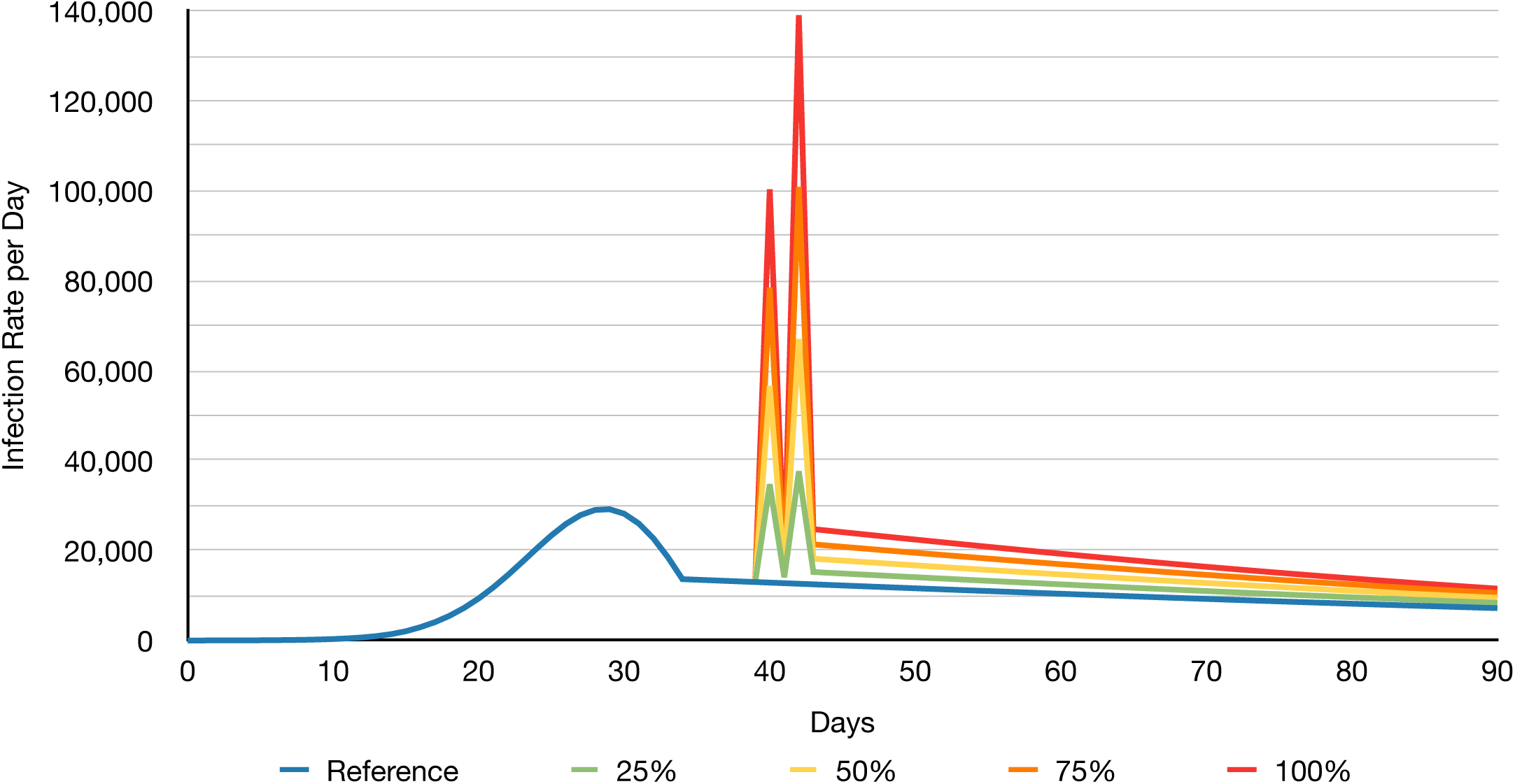
Scenario 3: Isolated Increases in Effective Contact Rate - Infection Rate showing the two peaks for the contact rate increases on day 40 and 42 with the latter ones being higher and amplified by the first one. Day 43 and later show the infection rates which are permanently increased due to the aftermath of the spikes prior to it.

The figures 6 through 9 on the next two pages demonstrate the effects that short outbursts can have and a few takeaways have to be mentioned and pointed out. First, a return to the effective contact rates of a “normal” state can increase the infection rates temporarily by 980% as the first day with increased effective contact rates amplifies the second one. This is due to the decrease in between those two dates not being sufficient for the measures to fight back the short upswing in a limited time. Therefore, these two increases could yield hundreds of thousands of new infections and thus could also even double the number of hospitalized patients. Second, in the long run, these short increases in effective contact rates can have detrimental impacts when it comes to the fatality numbers as a result of the increased hospitalizations. In the worst case, this could lead to an increase in fatality numbers of 60% after 90 days, not taking into consideration that hospitals might be overloaded and forced into triage procedure where limited resources have to be allocated and decisions have to be made which patients can receive treatments at all.

Overall, this scenario shows that singular increases already can have detrimental impacts and make the difference between hospitalization infrastructure being overloaded or able to handle the demand. In addition, the simulation shows the numbers immediately, whereas in reality, the incubation time might lead to a delay and thus the individuals infected over Easter could potentially affect the medical infrastructure one week to two weeks later.

With these aspects in mind, the last scenario will assess the worst possible option, a temporarily sustained increase, beginning on page 8.

### Scenario 4: Temporarily Sustained Increase in Effective Contact Rate

The above descried scenario assessed short singular increases, which might happen again in the future for certain events. This leaves the question though, since the last scenario already showed an interplay between two singular increases, how sustained increases, even if temporarily limited to various days, affect the numbers and if the reciprocity effects multiply. Therefore, this last scenario assesses a constant increase over Easter weekend, for example, if people would spend multiple days with family or other gathers, which is not unusual. Again, in order to simulate various severities of increases, four runs were conducted with differences of 25% yielding the past run as a return all the way to the effective contact rate of 10.325 for three days (Good Friday through Easter Sunday). The results are depicted in Figure 10 through 13 on the next pages and discussed thereinafter. Figure 14 shows the hospitalizations over 180 days for demonstration purposes.

**Figure 7.**
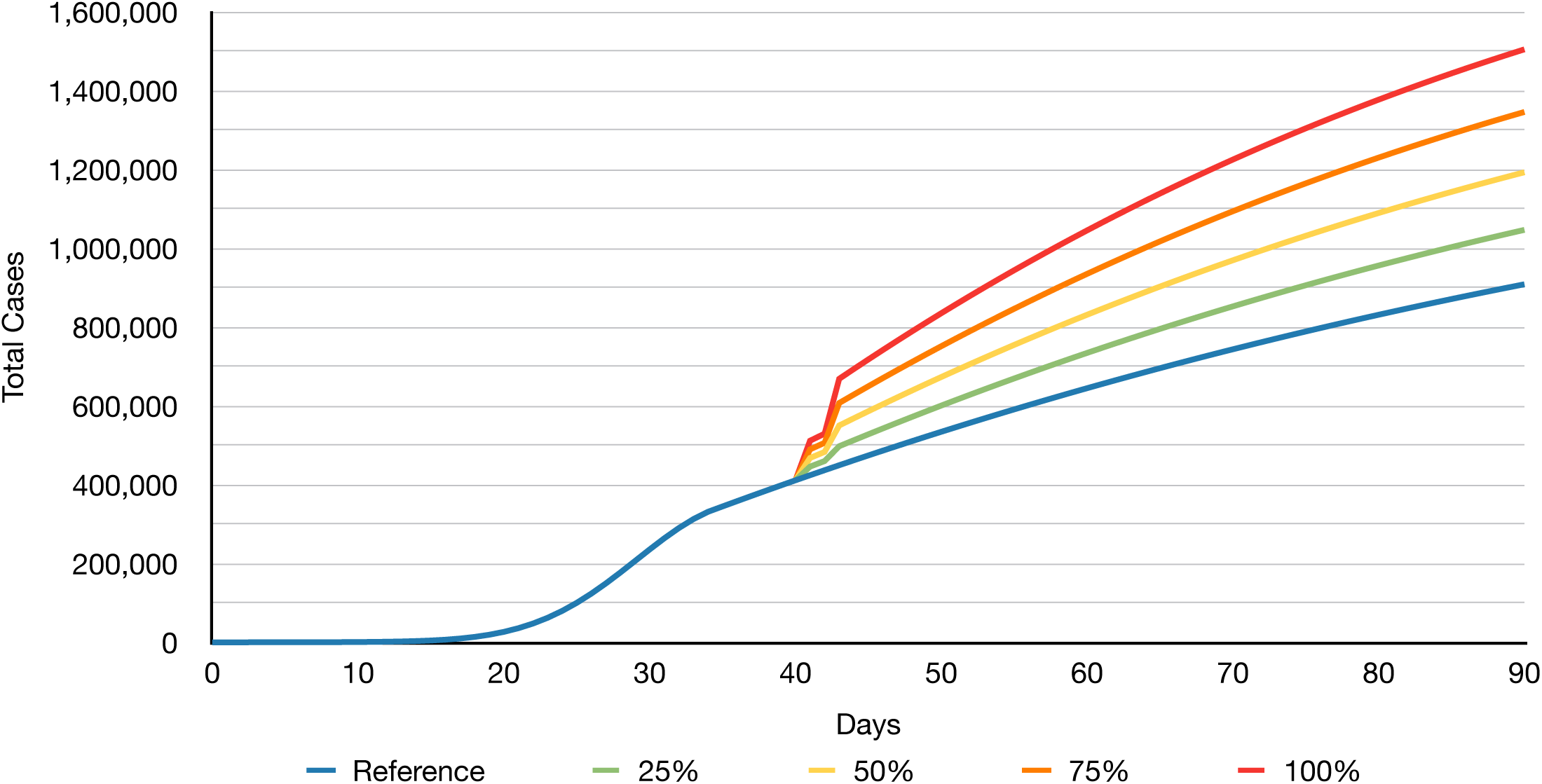
Scenario 3: Isolated Increases in Effective Contact Rate - Total Cases showing the effects of the two peaks on day 40 and 42 and the impact of the permanently increased infection rates over time in form of higher gradients for the simulated runs.

**Figure 8.**
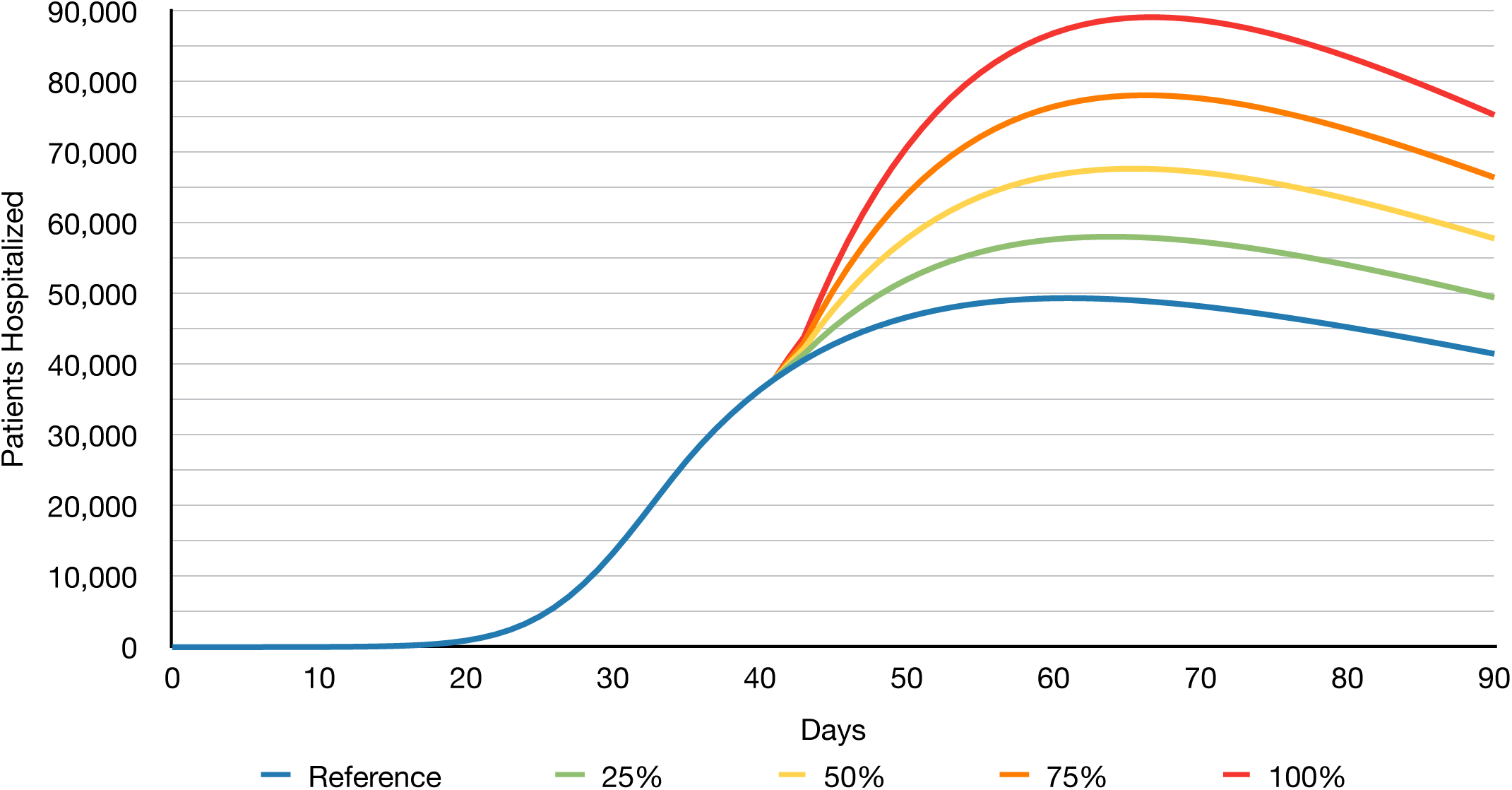
Scenario 3: Isolated Increases in Effective Contact Rate - Hospitalizations showing how many people require hospitalization for Scenario 3 each day after the delay of the incubation. This represents the required hospitalizations, which may exceed the real capacities of the hospitals and therefore cause shortage and possible even triage situations as described before. The predicted hospitalization numbers allow for estimation of necessary resources for the simulated area.

**Figure 9.**
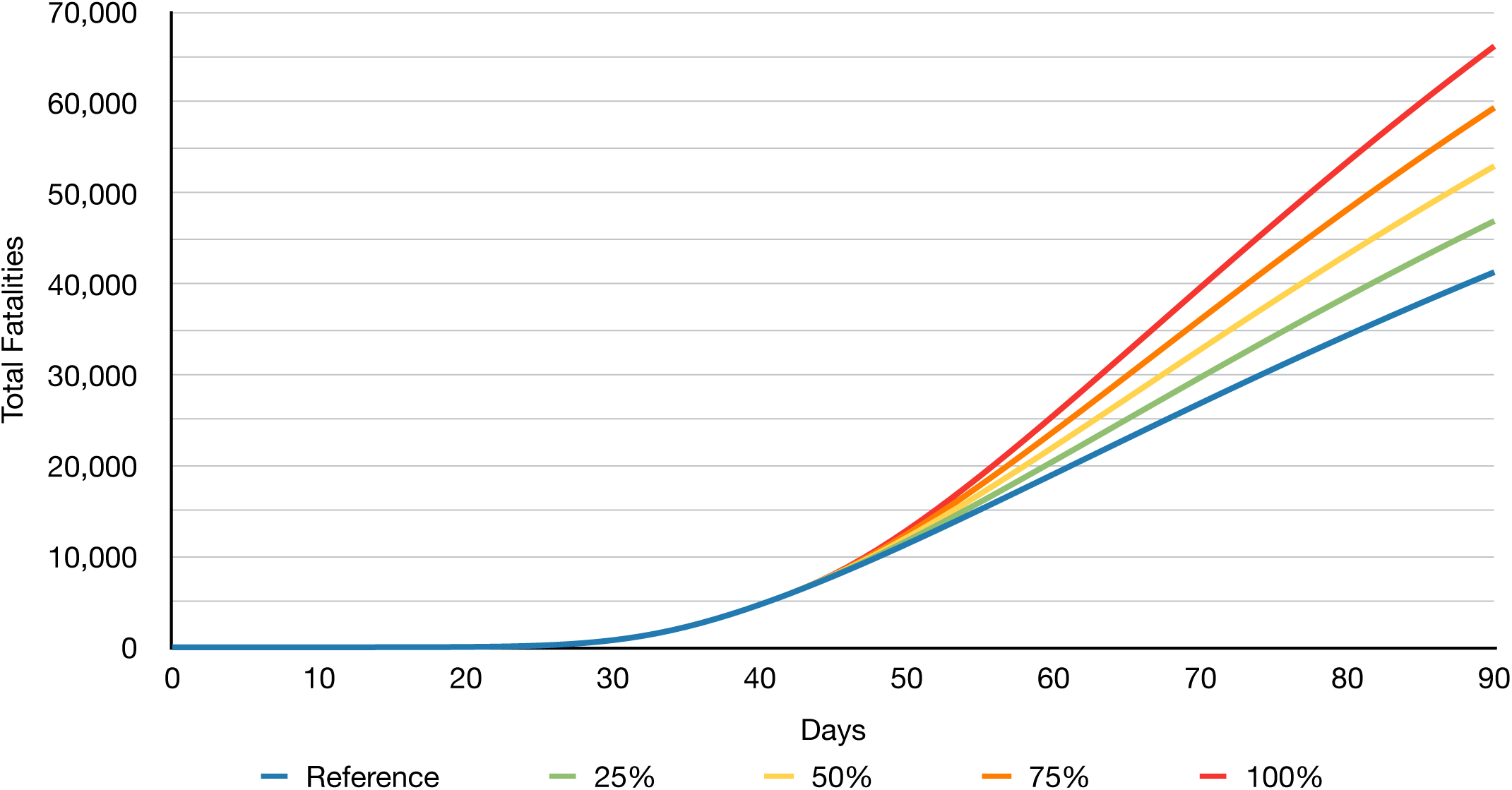
Scenario 3: Isolated Increases in Effective Contact Rate - Fatalities that show the increasing deaths over time with the different gradients based on the height of the peaks shown in the infection rates.

**Figure 10.**
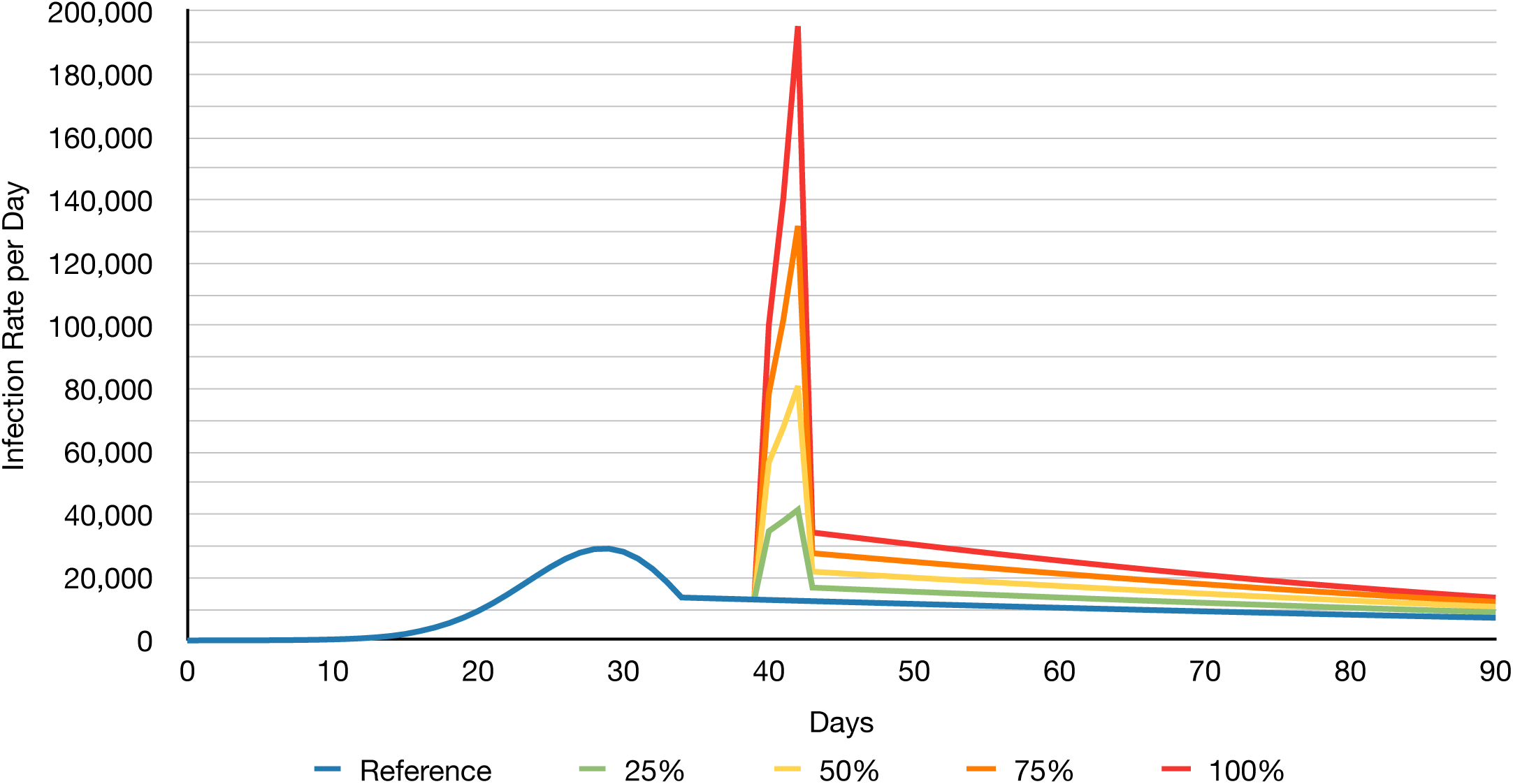
Scenario 4: Temporarily Sustained Increase in Effective Contact Rate - Infection Rate per day showing the constantly extremely increasing rates from day 40 through 42. These days each amplify the following one due to the sustained increased contact rates. This also exacerbates the effect after the subsidence of the peaks as the reaming infection rate is even more elevated that the one observed in Scenario 3.

**Figure 11.**
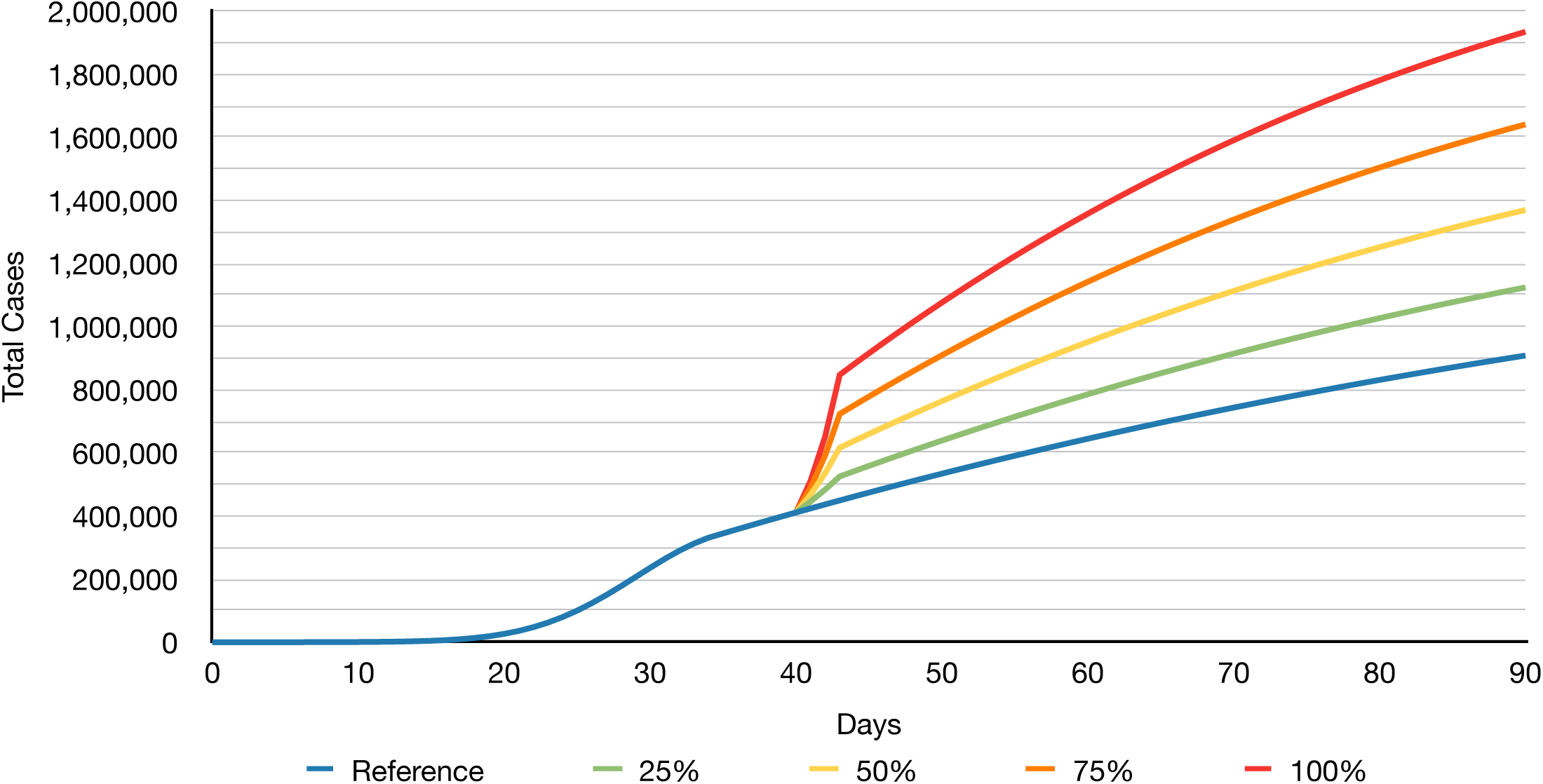
Scenario 4: Temporarily Sustained Increase in Effective Contact Rate - Total Cases showing the effects of the rapidly increasing infections from day 40 through 43 and the impact of the permanently increased infection rates over time in form of higher gradients for the simulated runs.

The figures resulting from the last scenario show that the effects are partially as to be expected based on Scenario 3 since the infection rate steadily rises with every day the increase persists and therefore the impact that the measures have when they are back in effect is also reduced. For example, for the infection rate on the first day after the increased period, the numbers are between 10% to 40% higher than they were in the respective runs of Scenario 3. This means that each day the increase persists will have permanent effects on the infection rates even once the effective contact rate goes back down. This permanent influence can have extreme ripple effects for the hospitalization and fatality numbers as shown by Figures 12 and 13 below: the hospitalization numbers are between 8.9% to 34.9% higher than the respective runs of Scenario 3 and between 29% and 146% higher than the reference run; the fatality numbers are between 6.8% to 28.6% higher than the respective runs of Scenario 3 and between 21.4% and 106% higher than the reference run over 90 days.

**Figure 12.**
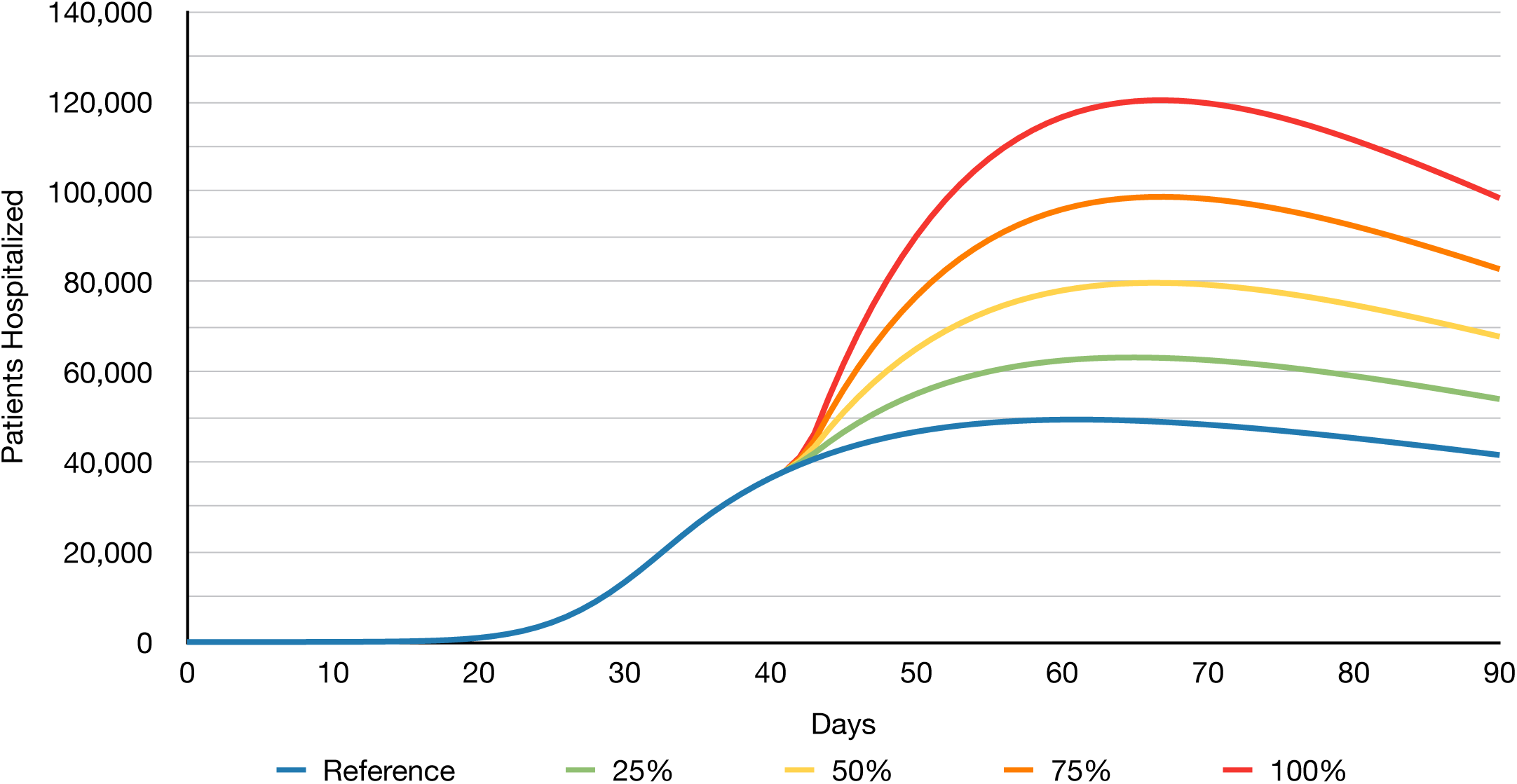
Scenario 4: Temporarily Sustained Increase in Effective Contact Rate - Hospitalizations showing even higher numbers of people requiring hospitalization after the delay of the incubation compared to Scenario 3. This results in even bigger loads for the hospitals and other infrastructure and presents an increased threat of collapse.

**Figure 13.**
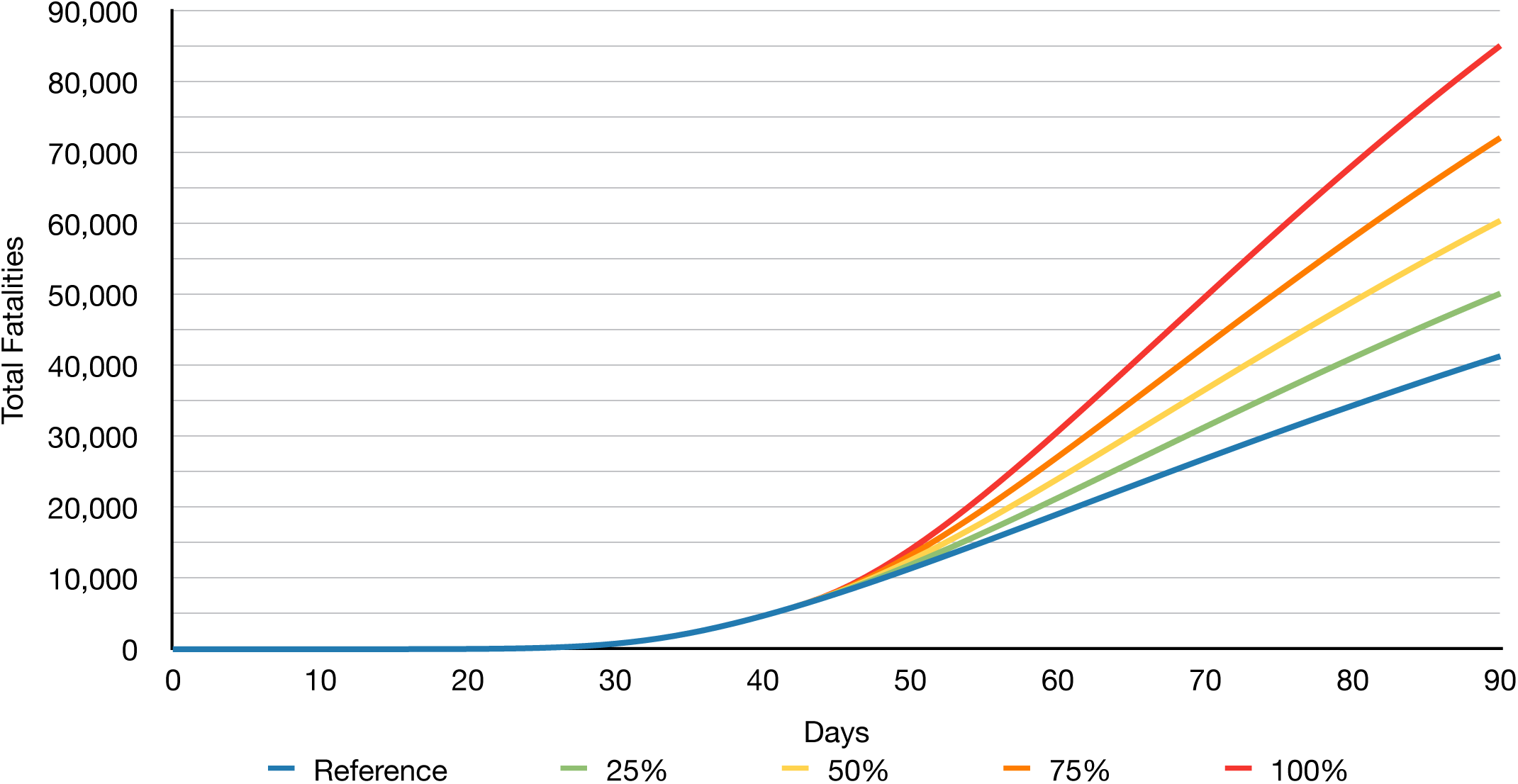
Scenario 4: Temporarily Sustained Increase in Effective Contact Rate - Fatalities over time showing the even higher numbers and gradients compared to Scenario 3 due to the sustained temporary increase in effective contact rates.

**Figure 14.**
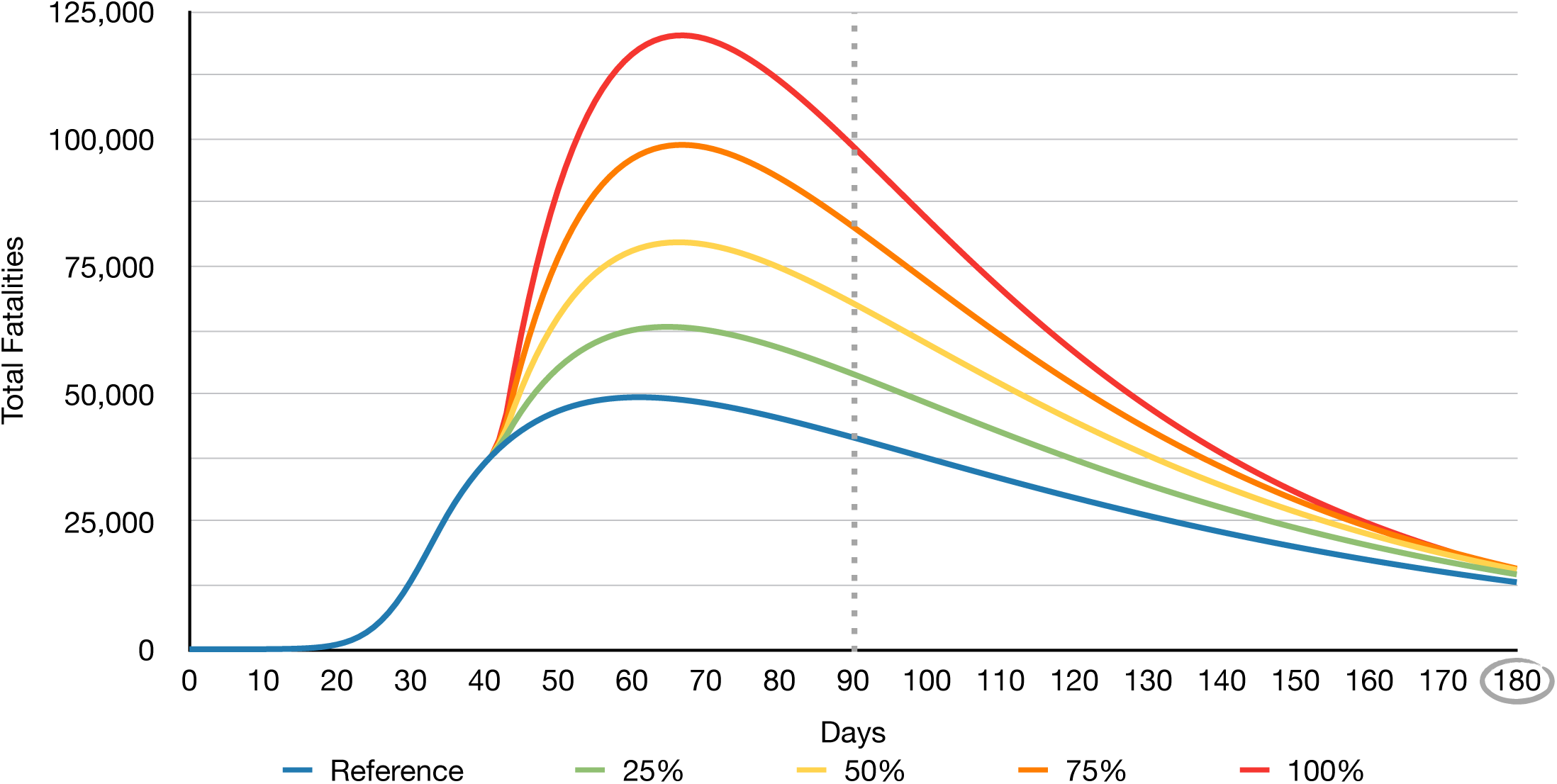
Scenario 4: Temporarily Sustained Increase in Effective Contact Rate - Hospitalization Rate over 180 Days showing the continued decline in required resources. This is dependent on a constant adherence to the measures and cannot be achieved with further deviations. It has to be noted though, that over the length of the simulation and with an increased time frame, the accuracy of the predictions declines due to possible unforeseen influences that are not included because they are not predictable.

All in all, we can see that a temporarily sustained increase not only increases the numbers and therefore causes effects over the time of its existence, it also affects the numbers after its subsidence as it permanently increases the severity of the pandemic. This allows for two conclusions: one, it is imperative to prevent such increases at any costs and two, if they are inevitable, they have to be kept as low and short as possible to minimize the permanent impact they have.

This concludes the simulations and scenarios assessed in this research. The last section will give a overview and summary including a conclusion and outlook regarding future research opportunities and plans.

## VI. Conclusion and Outlook

The previous sections have shown that the effects of such a dynamic and complex systems as this pandemic are by no means linear and predictable by mere extrapolation. Even with the measures and the current standings, short increase and maybe even returns to “normal” effective contact rates can have detrimental outcomes that cause permanent effects impossible to cure even when caught early. The two simulation scenarios 3 and 4 demonstrated that even single short increases can show these behaviors and temporarily sustained ones increase and amplify the impact through reciprocity.

Our simulations have shown that increases permanently increase infection rates after subsidence by as much as 40% and higher surges, such as a return to “normal” and therefore 100% increase of the effective contact rate would increase the infection rate temporarily by over 1,000%. These effects ripple through the system and impact hospitalizations and ultimately fatalities, increasing the former by as much as 146% at the peak and the latter by as much as 106% in the worst case compared to the references without contact increases.

In numbers, given that increases of 25% and 50% seem to be most likely given the data seen in Germany for the Easter weekend for example [2, 3], our simulations show the following increases (compared to realistic reference run) for a temporary 25% surge in contact rate: the total cases grew by 215,880, the maximum of required hospitalizations over time increased to 63,063, and the total climb in fatalities was 8,844 accumulated over 90 days. As for the 50% surge, we saw the total number of cases rise by 461,090, the maximum number of required hospitalizations increase to 79,733, and the total number of fatalities climb by 19,125 over 90 days in NYC.

In conclusion, the numbers and scenarios demonstrated that increases of any kind have to be prevent at any costs in order to not permanently impact the progress of the pandemic containment. If such increases cannot be prevented, it is imperative to keep them as short as possible and, if necessary, separate the peaks as much as possible in order to allow for regulation and mitigation in between. Furthermore, other mitigation strategies such as stricter regulations could be a possibility to mitigate already happened singular increases.

As described in the previous section, the results obtained in this simulation possess a certain predictive power within their numbers as they show possible phenomena, such as increases infection rates and their implications, before they develop effects in reality. This is especially important when it comes to the hospitalization rates, as increases infection rates or even short phenomena such as the described Easter Blip can significantly impact the hospitals in reality. Thus, the results allow a predictions to an extent when a wave as a result of an increase in infection numbers might come. This can allow authorities to assign resources accordingly or at least prepare for possible impacts especially since data seen in reality already shows the trajectory of the evaluated scenarios [2, 3].

As for future research and an outlook, other measures and effects, such as protective gear for the public can be assessed, as they might reduce the infectivity and or effective contact rate for example. This would allow for a selective use of such measures wherever necessary in order to purposefully utilize their effects. Moreover, other branches and population areas are planned to be researched, such as EMTs and police, as the impact of the pandemic on such forces is also important for the general public safety.

We see that the current pandemic impacts all our lives and will most likely continue to do so for, as of the time of this writing, an unexpected future. Fortunately the research conducted allows simulation and mimicking of the reality with predictive power and we will continue to adjust our models to include any new and important occurrences. Staying home and social distancing are our most powerful weapons in fighting this pandemic, but they only work if everyone participates, wide spread individual exceptions cannot be granted nor accepted and they can sabotage the whole mission. Let’s all do our part and participate in the fight, everyone can and everyone has to! Stay safe, stay home, stay healthy!

## Data Availability

Data and simulation models can be provided by the authors upon request. All simulation models and data sets are available and can be obtained if necessary.

